# Information Leaflets vs Artificial Intelligence: Comparing Perceptions of Stroke Survivors and Professionals in a Mixed-methods Study

**DOI:** 10.64898/2026.01.26.26344610

**Authors:** Lucie Tvrda, Jennifer K Burton, Katie McConnell, Kalliopi Mavromati, Hendrik Knoche, Robert Mikulik, Terence J Quinn

**Author notes:** Corresponding author: Lucie Tvrda.

## Abstract

**Background:** Stroke survivors often describe problems of insufficient access to information post-discharge. Traditional resources may not meet their information needs, but Artificial Intelligence (AI) could play a role.

**Aims:** To compare user perceptions of stroke information from third sector stroke websites with that generated by AI and summarize the attributes of the preferred stroke information formats.

**Methods:** UK third sector stroke websites were searched for materials relevant to 15 questions commonly asked by stroke survivors. ChatGPT(-4o) was used to generate responses to these questions. Stroke professionals (clinicians, researchers), stroke survivors, and their caregivers reviewed third sector and AI responses, indicating the source of the response and their preferred text. Participants also rated responses on scales of empathy, trustworthiness, reliability, comprehensibility and usefulness, and provided free text comments. Proportions of preference and correctly guessed responses, as well as mean ratings, were compared between the groups. Framework analysis was used to identify the attributes of response formats preferred by stroke survivors.

**Results:** Relevant responses were found for 13 (87%) out of 15 questions. Across groups, 60 participants with mean age of 44 (SD=14) and 57% females, correctly identified 184/300(61%) of AI responses, and preferred AI response in 123/300(46%) of the cases. The groups differed in their preference with clinicians being least likely to choose AI (34%), followed by stroke survivors (49%) and researchers (54%). All groups viewed third sector responses as more empathetic in tone. The themes of content, structure, and tone of responses were described by stroke survivors with the emphasis on clarity, conciseness, and approachable tone.

**Conclusions:** AI-generated responses to stroke questions were rated positively by stroke survivors and researchers, whereas stroke clinicians were more sceptical. Given that stroke information materials are intended for people with lived experience of stroke, their input should be prioritized to inform development of new resources.

## Introduction

Timely and reliable information is essential to support informed decision-making after stroke, and improve long-term stroke outcomes[1]. Stroke survivors and their families frequently report limited access to information following discharge from hospital[2]. Information delivered by healthcare professionals is preferred, however, inadequate and insufficient communication may lead to unmet information needs[3].

Third-sector services offer stroke information leaflets created by healthcare and other professionals. The American Medical Association (AMA) recommends that healthcare materials are produced at a sixth grade reading level to increase the accessibility of health education[4]. While a helpful resource, current stroke information leaflets often exceed this level, rendering the information difficult to understand[5]. This impacts stroke survivors’ health literacy which may lead to poorer health outcomes[5]

Large language models (LLMs) are increasingly used in healthcare due to their ability to mimic natural language[6]. An LLM flagship, ChatGPT, has performed well in producing clear and concise medical summaries[7]. ChatGPT can also increase comprehensibility by adjusting the text to the literacy level of a reader living with a medical condition[8]. However, clinicians have criticized the accuracy of Artificial Intelligence (AI)-generated medical information[9]. LLMs may not be trained on trusted medical data and provide unreliable information while appearing confident, a phenomenon known as “hallucinations”[10]. This, in turn, may harm the level of human trust, and poses risks if used to inform medical decisions[11].

Studies have found that the general population perceives AI-generated medical advice as less empathetic and reliable than that written by a human[12]. Conversely, people living with a medical condition thought AI was able to provide a more empathetic response than a healthcare professional[13]. Stroke survivors have reported perceived benefits of AI and digital tools in stroke care (e.g., digital home rehabilitation)[14], but their perceptions of AI as a source of stroke information remain unknown.

This study had three aims:

1. To identify the most common questions following stroke and evaluate the availability of responses found at online third sector stroke websites across the UK.
2. To compare the perceptions of stroke survivors, researchers and stroke clinicians rating information materials produced by third sector stroke websites, and those generated by ChatGPT 4.o.
3. To summarize the characteristics of stroke responses across common topics, in a way that meets the needs of people with lived experience of stroke.

## Method

### Ethics and Recruitment

This mixed-methods study followed the Strengthening the Reporting of Observational Studies in Epidemiology (STROBE) and the Standards for reporting qualitative research (SRQR) reporting guidelines (Supplementary) and was approved by the University of Glasgow College of Medical, Veterinary and Life Sciences Ethics committee (200220163). Written informed consent was obtained from all participants.

Participants were recruited by convenience sampling, using the NHS Research Scotland Stroke User Group of stroke survivors and professionals.

We recruited adults who self-identified as: (1) stroke survivor, carer or family member of a stroke survivor; (2) stroke researcher; or (3) stroke healthcare professional. We aimed for inclusive recruitment with no exclusions around stroke related impairments. Participants with no access to digital technology were offered in-person interviews.

### Materials and Procedure

#### Common Stroke Questions

We compiled a list of commonly asked stroke questions, drawing together interviews with stroke survivors and their families[15] and the UK Stroke Association helpline log, listing the most frequent enquiries to this telephone advice resource between the years 2023 and 2024 (Supplementary Table 1). We identified 15 questions, i.e., the three most common questions from each of the five overarching topics: general information, health issues, stroke recovery, life after stroke, support after stroke[3].

#### Stroke Responses

We mapped the availability of responses to each of our 15 questions on four UK third-sector websites (Stroke Association, Northern Ireland Chest Heart & Stroke, Different Strokes, Chest Heart & Stroke Scotland), using a classification of: not available; not directly available but related information may be found; available. The online resources we used had not been produced using AI.

Two stroke researchers (LT and KMC) then compiled responses to the 15 questions from online materials. Where more than one website provided relevant information, we selected one that best covered the question based on agreement between the researchers. Additionally, we used ChatGPT 4.o to generate a response to each of the 15 questions (e.g., what are the side effects of different stroke medications?), limiting the length to 300 words with no additional prompts. ChatGPT was chosen due to its superior readability of patient information compared to other LLMs[16]. Any information that could potentially identify the source was removed.

#### Measures

We collected demographic measures including age, gender, and participant group (healthcare professional; researcher; stroke survivor or family member). We assessed attitudes towards AI using the 5-item Likert AI Attitude Scale (AIAS-4)[17]. We further assessed perceptions of responses: reliability, empathy, comprehensibility to someone who had a stroke, and willingness to use the information, through 5-point Likert scales ranging from a very negative to a very positive perception (e.g., ‘very unreliable’, ‘somewhat unreliable’, ‘neutral’, ‘somewhat reliable’, ‘very reliable’)[12]. The fifth scale, relevance, was added following the same format, to evaluate how well the materials answered the question. Clinicians further rated each response on 6-point Likert scales of accuracy and completeness[18].

#### Procedure

Semi-structured interviews were conducted by LT between October 2024 and May 2025, in person or online using Microsoft Teams(v.25241.203.3947.4411.). Interviews were scheduled to last no more than one hour with breaks to prevent fatigue. The first 5 clinician interviews were considered pilot to refine the testing session delivery and included in the dataset if no major changes were required. Participants rated two responses (third-sector website and AI) to five randomly selected questions, one for each topic, blinded to the source. The two responses were presented sequentially in a random order allocated using RStudio (v.4.1.2)[19], to minimise response bias. Enough time was given to the participant to read and rate the response on each of the rating scales.

Participants were asked to guess the source of the response (third-sector website or AI) and indicate their preference, briefly exploring the reasoning behind their choice. This provided a valuable source to inform the development of future materials addressing unmet needs. If the participant’s preference matched their guess this was treated as a “conscious preference” (e.g., guessing AI as the source, and preferring the AI response). If consented by the participant, the interviews were audio-recorded and verbatim transcripts produced. All transcripts were anonymised prior to data extraction.

### Quantitative Analysis

Power analysis was conducted using the G*Power tool (v.3.1.9.7.) to identify the minimum sample size required to detect a moderate effect size on an alpha level of 0.05. Quantitative data were tabulated using Microsoft Excel (v.2509). Analyses and visualisations were conducted in RStudio (v.4.1.2)[19]. We used descriptive statistics to assess the distribution of participants’ guesses, preferences, and ratings. All missing values were excluded prior to the analysis. Chi square test was used to compare correct guesses, preferences, and conscious preferences across the three groups. Paired t-tests were used to evaluate the differences in ratings between groups.

### Qualitative Analysis

All transcripts were imported to NVivo (v.14)[20]. We used the framework analysis approach[21] with the *a priori* interest to identify the attributes of responses favoured by stroke survivors and their families.

LT initially screened all transcripts to build a foundation for the analytical framework. Any recurrent points and descriptions of preferred responses were noted.

A random sample of four transcripts were initially coded line-by-line by LT to identify expressions by which quality of a stroke response was characterised. These transcripts were also coded by co-investigators (LT, KM, JB) to capture a wider range of codes and categories (i.e., potential response attributes).

After the initial codes were compared, we created a set of codes which were grouped into categories forming the analytical framework. This was applied to the remaining transcripts and developed iteratively, gaining its final shape after the last transcript has been coded and indexed.

We constructed a framework matrix for each of the five question topics. Each row represented a participant, with a column for each final theme (response attribute). Individual quotations were imported into the matrix (Supplementary Tables 2-6).

Shared patterns of meaning identified from the framework matrices were discussed within the team and summaries of commonalities and differences of preferred response attributes were described by topic, as themes (Supplementary Table 7).

## Results

### Common Stroke Questions

Previous interviews with stroke survivors and family members (N=32 stroke survivors, 2 caregivers), and Stroke Association helpline log search revealed the 15 most common stroke questions following discharge from hospital (Table 1), representing 40% of all the 27,439 helpline enquiries. Relevant online responses were found for 13(87%) of the questions. There was no dedicated online information resource for two questions: *What are the side effects of stroke medications?* and *Is stroke a genetic condition?*. Of the four online sources Stroke Association had the highest number of responses available.

**Table 1:**
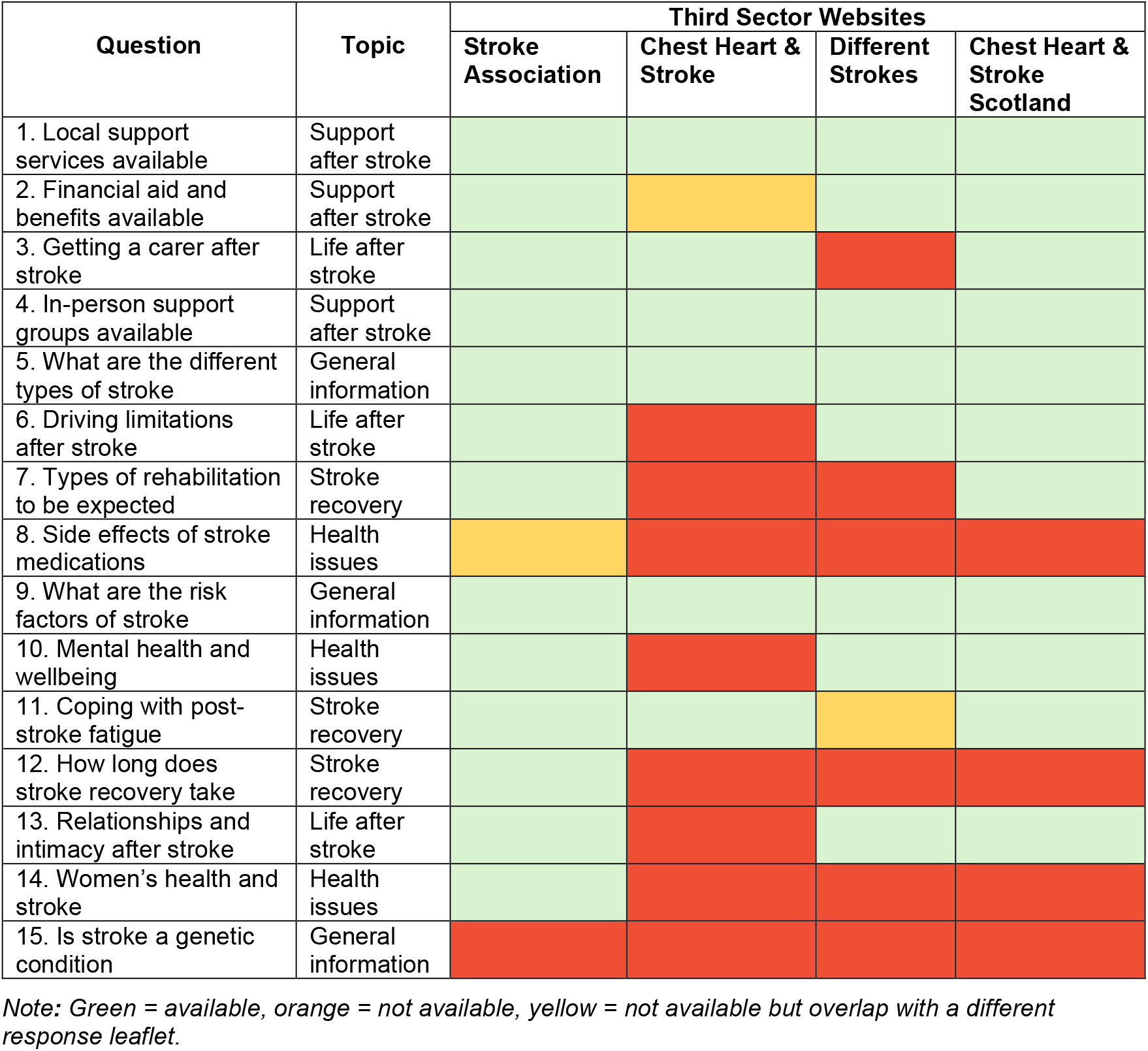
Overview of information leaflets available on UK third sector stroke websites.

### Information Source Preference

We recruited 20 stroke healthcare professionals, 20 stroke researchers, 17 stroke survivors and 3 caregivers/family members. In total, 60 participants (57% female) with mean age of 44(SD=14) compared third-sector website and AI responses (Table 2). Four interviews were conducted in-person. The 5(8.3%) pilot participants did not report their age, otherwise there were no missing values. Preferences differed significantly across groups (*X*^2^(2)=8.7, *p*<.05) with clinicians being least likely to select AI as their preferred response.

**Table 2:** Descriptive and comparative statistics.

|  | Clinicians | Researchers | Stroke survivors | Comparative statistic |
| --- | --- | --- | --- | --- |
| N | 20 | 20 | 20 |  |
| Age (M, SD) | 43.4 (11.1) | 32.3 (9.6) | 55.3 (12.1) |  |
| AI Attitude (M, SD) | 7.66 (1.15) | 7.62 (0.91) | 5.72 (2.34) |  |
| Guessed AI correctly (N, %) | 66/100 (66%) | 65/100 (65%) | 53/100 (53%) | $p=.110$ |
| Preferred AI response (N, %) | 34/100 (34%) | 54/100 (54%) | 49/100 (49%) | $p=.012$ |
| Consciously preferred AI (N, %) | 8/100 (8%) | 29/100 (29%) | 17/100 (17%) | $p<.001$ |

*Note: N=number; M=mean; SD=standard deviation; %=percentage; AI=artificial intelligence; p=p value; higher values on the AI Attitude scale represent a more favorable approach towards AI.*

When grouped by question topic, participant preferences were varied with significantly higher proportions of clinicians preferring website information for topics of health issues, stroke recovery and support after stroke. AI responses were favoured by researchers for topics of life after stroke and general stroke information (Figure 1). Stroke survivors did not have a significant preference in any topic.

**Figure 1:**
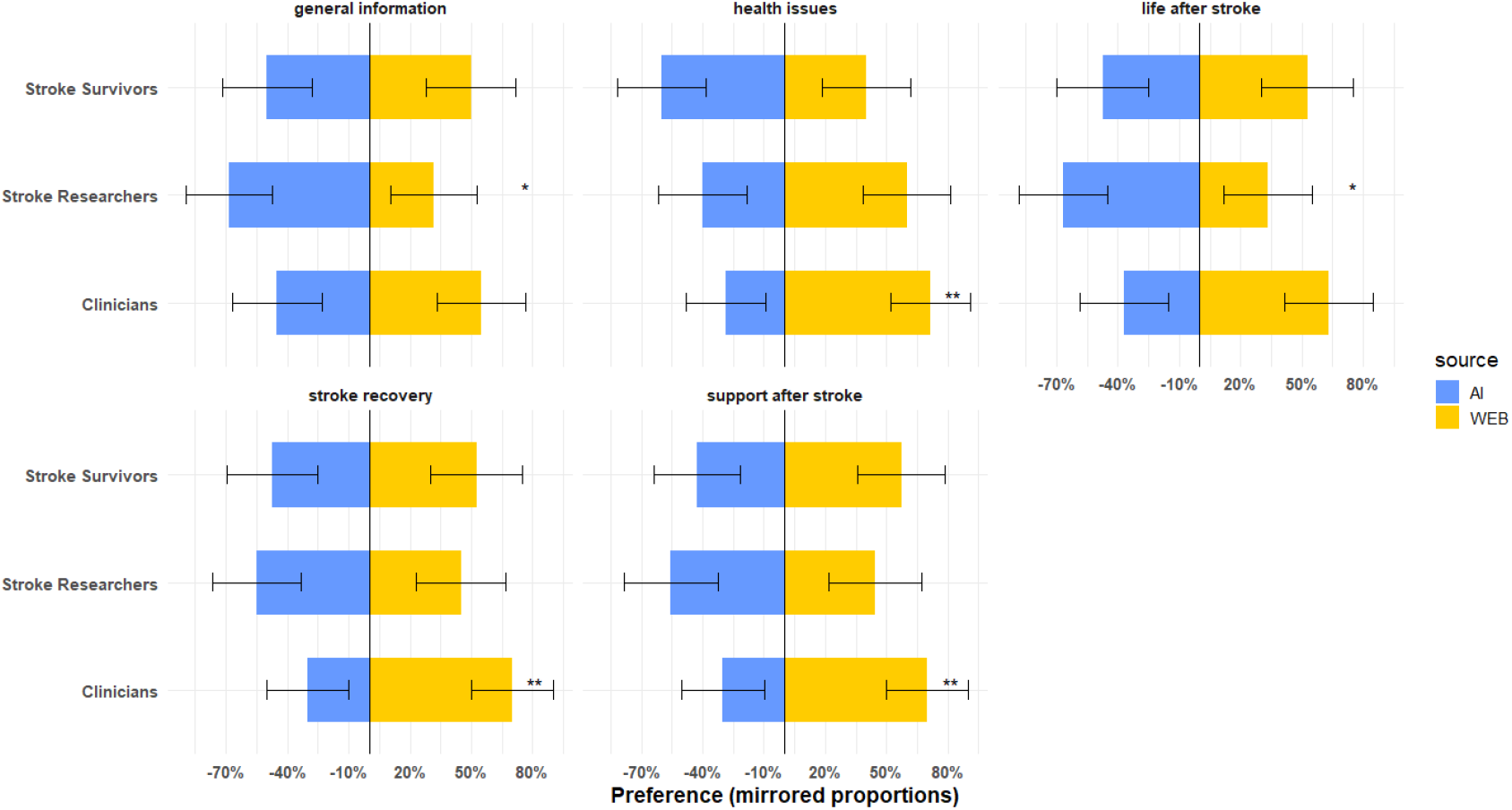
Bar plots representing participant preferences across stroke topics. Note: Proportion of participants preferring a response by artificial intelligence (AI) or third sector website (WEB) across stroke topics.%=percent; * = p<.05, ** = p<.01; the error bars represent 95% confidence intervals.

### Ratings of Stroke Responses

Third-sector responses were rated as significantly more empathetic than AI by clinicians (*p*<.001) and researchers (*p*<.05). Additionally, clinicians were more willing to use third-sector leaflets (p<.05) and rated them as more comprehensible (*p*<.05). Stroke survivors’ ratings did not differ between AI and third-sector responses (Figure 2). Out of 100 rating instances, clinicians found 33% AI responses and 40% of third sector materials completely accurate. Considering completeness of the information, clinicians rated 32% AI responses and 23% third sector responses as complete. These differences were not significant.

**Figure 2:**
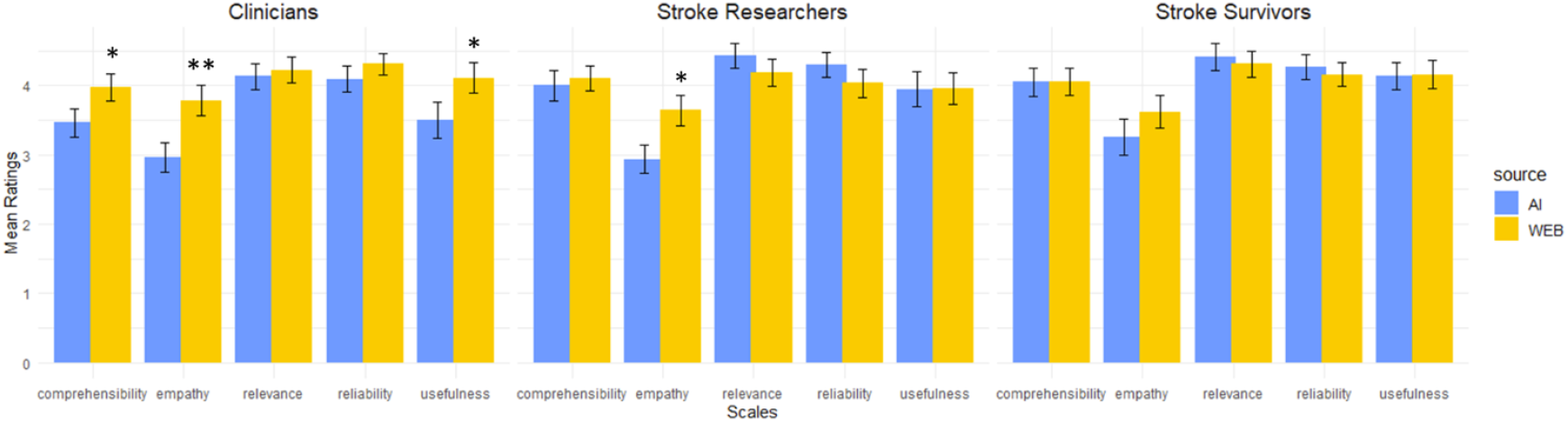
Bar plots showing mean ratings across participant groups. Note: Mean ratings of Artificial Intelligence (AI) and third sector website (WEB) materials on 5-point Likert scales of empathy, comprehensibility, usefulness, reliability, and relevance. Higher number corresponds to a more favourable rating. * = p<.05, ** = p<.01.

### Qualitative results

As two testing sessions were not recorded, eighteen semi-structured interviews with 15 stroke survivors and 3 caregivers/family members were included in the qualitative analysis. Three themes were generated, reflecting preferred response attributes: content, structure and tone (Table 3). The three attributes were evaluated across five question topics (general information, health issues, stroke recovery, life after stroke and support after stroke)[3].

**Table 3:** Framework analysis themes illustrated by quotations.

| Quotation No. | Participant characteristics | Quotation |
| --- | --- | --- |
| **Content** |  |  |
| 1 | male, 51-55 | *"Response A was really straightforward and down the line and B seemed to sort of meander around a bit more, was a bit more vague. But I definitely like A's presentation more."* |
| 2 | male, 56-60 | *"It just kind of said things, said facts, words. It didn't put any context or meaning to them."* |
| 3 | male, 51-55 | *"It was lots of information. You might not need all the information."* |
| 4 | female, 21-25 | *"It lists all the different places which I don't know if I did like. So, it's that I almost didn't trust that it was giving specifics anyway."* |
| **Structure** |  |  |
| 5 | female, 46-50 | *"(...) waffly, too much information. It wasn't relevant (...) and the second one was very clear and concise."* |
| 6 | male, 71-75 | *"[...] This is a bit easier to understand, especially for someone like me. (...) my memory is - I'm not as fast, I'm not as reliable reading paragraphs like that"* |
| 7 | male, 46-50 | *"(...) so it just gives you a list of all the things which I think would be easier for people to understand."* |
| 8 | male, 76-80 | *"(...) it was more sort of strict bullet points of boom, boom, boom."* |
| **Tone** |  |  |
| 9 | female, 56-60 | *"Very clinical, not any personable information about it[...]. I'm looking for something, maybe a wee bit of sugar-coated."* |
| 10 | male, 56-60 | *"(...) it was more emotional. Mostly, the first one just seemed to give you the steps you would have to follow. The second one seemed to be a wee bit more empathetic."* |
| 11 | female, 71-75 | *"I felt it was a bit schoolteacher-ish. You know, you must do this and this because it's going to make you part of the community [...] What if you don't want a sense of community?"* |
| 12 | female, 56-60 | *"After having a stroke, the biggest feeling is fear - fear of what's coming. And then it never said that it would be hard to go to these groups. I found that very hard."* |

*Note: Participant characteristics include gender and age group in years.*

#### Theme 1: Content

In general, stroke survivors thought that information resources should be focused and directly address the question while avoiding irrelevant tangents (quotation 1). Within the individual topics, detailed explanations providing richer and more meaningful understanding of the information were desired when talking about stroke-related health issues. For instance, one stroke survivor disliked a website leaflet addressing post-stroke health plainly (quotation 2). Similarly, participants felt that responses to questions about stroke recovery should contain specific actionable advice, information with examples, and clear explanations of medical procedures and terms. In contrast, the preferred format of information relating to life after stroke was short and focused (quotation 3). Participants expressed varied needs for information about stroke support. Some preferred a generalised response while others needed a targeted personalised response. However, all agreed that very specific information came across as irrelevant and unreliable (quotation 4).

#### Theme 2: Structure

Ease of understanding was perceived as the most desirable quality of the stroke information structure across all question topics. Participants agreed that clear and concise responses with short sentences were preferrable (quotation 5), which was also mentioned in the context of post-stroke fatigue and cognitive problems (quotation 6).

The five topics differed slightly in the preferred information format, with a bullet-point list being favoured for general information about stroke (quotation 7). Conversely, when talking about stroke recovery, while a clear and concise response was still desired, participants found bullet-points overly rigid (quotation 8).

#### Theme 3: Tone

Overall, stroke survivors thought that stroke information materials should be written in plain language with an approachable, gentle tone. Excessive use of medical terminology was perceived negatively, with the term “scary” specifically used. Very direct responses were also not favoured (quotation 9).

Within individual question topics, participants preferred warm, empathetic, and emotional responses to questions about life and recovery after stroke, as well as health issues (quotation 10). Conversely, emotional tone in leaflets providing practical information about post-stroke support was perceived as patronising by some (quotation 11). The failure to acknowledge barriers to accessing the support available was also noted (quotation 12).

## Discussion

We have shown that current UK online stroke information materials do not cover all common post-discharge questions, and that stroke interest-holder groups differ in their perceptions of AI as a source of stroke information. We have highlighted the necessity for materials that are concise, approachable and easy to understand, but also demonstrated differing format and content preferences among stroke survivors, according to the subject matter.

Most healthcare professionals preferred third-sector resources whereas researchers were more inclined towards AI-based materials, and the preferences of stroke survivors were evenly split. These findings are in line with evidence from other healthcare areas showing that clinicians are more critical towards AI-generated content than people living with relevant health conditions, rating it as less understandable[9, 22], and less accurate[9]. Interestingly, clinicians in our study held the most positive general attitudes towards AI, as rated on the AI Attitude scale, suggesting that the true differences in perceptions of AI in healthcare are dependent on professional role and experience rather than age.

Clinicians and researchers rated the third-sector leaflets as significantly more empathetic than AI, which is in agreement with previous research where people’s perception of an empathetic medical response emerged from knowing that the text was human-written[12]. In our study, participants were masked to the source of the response to avoid bias. We have thus demonstrated that even if unaware of the authorship, stroke professionals perceive human-written content as more empathetic. However, an unconscious negative bias towards AI was present, as participants were likely to prefer responses, which they labelled as human-written even if they were, in fact, AI-generated.

In contrast, stroke survivors’ empathy ratings did not differ between AI and third-sector leaflets. Nevertheless, while expressing a desire for empathetic and emotional tone in health-oriented materials, more than half of stroke survivors favoured AI-generated responses to questions about health issues. In previous research, cancer patients rated chatbot responses as more empathetic than clinician responses[13]. This suggest that healthcare professionals’ understanding of empathy may not be fully aligned with the needs of people living with a medical condition[23]. The importance of empathetic communication in healthcare settings is known with studies emphasizing its positive effects on treatment adherence and recovery[24]. Notably, stroke survivors felt that the emotional tone should be used with caution when providing facts and practical information. Responses that motivate and suggest options while supporting personal choice were favoured. This supports previous evidence highlighting the need for autonomy among stroke survivors[25].

Reasoning provided by the stroke survivors and their families enabled us to make recommendations for the content, structure and tone of stroke information materials. Specifically, responses to questions about health issues and recovery should be written in conversational style, contain actionable advice and explain medical facts with empathy, from the patient’s perspective. In contrast, general stroke information may be provided as a bullet-point list and stay factual, with a country-specific focus. We recommend that all information materials are clearly structured and concise using short sentences that are easy to read. We note that this is aligned with the characteristics of a sixth-grade reading level recommended by AMA as the basis for all patient-facing healthcare materials[4].

### Strengths and Limitations

The mixed-method approach allowed us to compare perceptions across groups while also gaining a deeper understanding of the stroke survivors’ information needs. Moreover, the involvement of stroke researchers allowed us to understand the perspectives of a group often overlooked in literature. Due to the hybrid nature of this study, we were able to recruit a wider range of participants including those with limited access to technology. Our qualitative analysis was purposively focused to make recommendations informed by the participants’ insights.

Despite the inclusive measures, this study was limited by a potential selection bias with a tendency for the participating stroke survivors to be younger than the general stroke population[26]. As we have only included frequently asked questions, we did not evaluate the ability of AI to respond to less common enquiries, which are more prone to hallucinations[10]. Furthermore, the response rating may have been biased by the fatigue effect due to the length of the testing sessions. Although, we aimed to minimise length and offered comfort breaks. The present sample size is a limitation in that we could not control for confounding factors, such as age, gender and AI attitudes in a robust manner. Lastly, the present results may not generalise to other countries, as only UK websites were included.

### Implications and Future Directions

This study has shown that currently available stroke information is incomplete, and the information needs of people affected by stroke are not fully aligned with opinions of clinicians and researchers. AI-generated responses were well received by stroke survivors, suggesting AI’s potential to complement existing stroke information resources. These findings highlight the importance of user involvement in the development of stroke information materials to maximise accessibility and effectiveness. The current recommendations for response formulations may also aid with generating prompts to tailor AI responses and with designing digital assistive technologies using LLMs to address stroke-related questions.

Future studies may further assess perceptions of stroke information formulations using the present recommendations. A larger sample size for the quantitative analysis would enable robust cross-group, and potentially, cross-country comparisons.

## Conclusions

In this mixed-method study, we have shown that current third sector information materials did not cover all common questions after stroke, and many were not accessible to stroke survivors. AI-generated responses to stroke questions were perceived well by stroke survivors and researchers, while clinicians remained sceptical. To increase accessibility of the information, stroke responses should be clear and concise, while avoiding excessive medical terminology. Warm and empathetic responses are needed to answer health-oriented questions. We emphasise the importance of incorporating the input from people with lived experience of stroke in the development of future information resources.

## Statements and Declarations

## Acknowledgements

We thank the Registry of Stroke Care Quality Plus (RES-Q+) consortium for providing support and guidance in the conception and dissemination of this research. We also thank the NHS Research Scotland Stroke User Group team for their support with participant recruitment.

## Author contributions

LT: conceptualization; investigation; analysis; visualization; writing original manuscript; review and editing. JB: conceptualization; validation; review and editing. KMC: resources; investigation; review and editing. KM: validation; review and editing. HK: supervision; review and editing. RM: funding acquisition; review and editing. TQ: project administration; conceptualization; resources; supervision; review and editing.

## Ethical considerations

This study was approved by the University of Glasgow College of Medical, Veterinary and Life Sciences Ethics committee (200220163).

## Consent to participate

Written informed consent was obtained from all participants.

## Consent for publication

Not applicable.

## Conflicts of interest

The authors have no conflicts of interest to declare.

## Funding

This study was funded by the European Union (EU) as part of the Horizon Europe research initiative RES-Q+ (grant number 101057603). Views and opinions expressed are, however, those of the authors only and do not necessarily reflect those of the EU or the Health and Digital Executive Agency. Neither the EU nor the granting authority can be held responsible for them.

## Data availability

Data supporting the present findings are available upon reasonable request.

## Notes

### Competing Interest Statement

The authors have declared no competing interest.

