## Supplemental Materials for "Information Leaflets vs Artificial Intelligence: Comparing Perceptions of Stroke Survivors and Professionals in a Mixed-methods Study"

**Table 1: List of questions commonly asked by stroke survivors after being discharged from hospital.**

| **Number** | **Question** |
| --- | --- |
| 1 | What are the local support services available after stroke? |
| 2 | What financial aid and benefits is available after stroke? |
| 3 | How can I get a carer after stroke? |
| 4 | What in-person support groups are available in my area? |
| 5 | What are the different types of stroke? |
| 6 | What are the driving limitations after stroke? |
| 7 | What types of rehabilitation can be expected after having a stroke? |
| 8 | What are the side effects of stroke medications? |
| 9 | What are the risk factors of stroke? |
| 10 | How does stroke impact mental health? |
| 11 | How to cope with post-stroke fatigue? |
| 12 | How long does recovery take and how to know if I will make full recovery? |
| 13 | How does stroke affect relationships and intimacy? |
| 14 | How does stroke affect women? |
| 15 | Is stroke a genetic condition? Did I inherit it? |
| 16 | Why did my stroke happen? |
| 17 | Can stroke lead to dementia? |
| 18 | Can stroke lead to depression? |
| 19 | What are statins for? |
| 20 | Could my past stroke have been prevented? |
| 21 | Can I prevent having another stroke in future? |
| 22 | Will my visual problems recede? |
| 23 | Why might additional brain imaging be needed? |
| 24 | What additional tests may I need? |
| 25 | What are the odds that I will have another stroke? |
| 26 | Does diabetes increase my risk of having a stroke? |
| 27 | Do my cholesterol levels put me at risk for a stroke? |
| 28 | Is my weight within a healthy range to prevent a stroke? |
| 29 | Is my blood pressure within the normal range? Can you help me control it? |
| 30 | Does having a stroke increase my risk of having a heart attack? |
| 31 | When are blood thinners necessary? How is this decision determined? |
| 32 | What are the differences between blood thinning medications? |
| 33 | What are the risks of blood thinning medications? |
| 34 | What are the side effects of different stroke medications? |
| 35 | What percentage of stroke survivors are independent in daily living? |
| 36 | When can I go back to work after stroke? |
| 37 | Can you help me quit smoking? |
| 38 | Do I need to implement a special diet after stroke? |
| 39 | Why do I feel so overwhelmed and fatigued? |
| 40 | Will I be able to travel and go on holidays? |
| 41 | Will I be able to use public transport? |
| 42 | Will I be able to dress myself properly? |
| 43 | Will it be hard to get back to running/being aerobically fit again? |
| 44 | Will this affect how long I have to live? |
| 45 | Will I be able to wear high heels again? |
| 46 | Will I be able to do everyday manual tasks (using a knife and fork, opening jars)? |
| 47 | Can I, or will I be able to enjoy the quality of life I had before the stroke? |
| 48 | Will I ever return to my pre-stroke situation? |
| 49 | Will I be able to regain my flexibility, balance & endurance? |
| 50 | Will I be able to coordinate my body movement better? |
| 51 | At what point will my movement start to come back in my hand/foot? |
| 52 | Will I be able to walk normally again after the stroke? |
| 53 | Will I be able to write again? |
| 54 | Will I be able to speak and understand conversation properly again? |
| 55 | How much rehabilitation will I need? |
| 56 | What is the timing, intensity, and duration of physiotherapy activities I need to do? |
| 57 | How much do I need to rely on my own motivation to recover? |
| 58 | Does the type, severity and site of my stroke impact my recovery potential? |
| 59 | Will my confidence return? |
| 60 | Is it true that there is a cut-off point for functional recovery? |
| 61 | Will I be able to regain a high degree of independence? |
| 62 | For how long will I receive medication? |
| 63 | What combination of lifestyle, medication, and in-hospital treatments/surgery/rehabilitation may be necessary? |
| 64 | What are the likely stroke recovery outcomes? |
| 65 | What happens after treatment? How long will recovery take? |
| 66 | What kind of rehabilitation should be expected after having a stroke? |
| 67 | What can you tell me about stroke? |
| 68 | What are the things that can help prevent a stroke? |
| 69 | What are the risk factors associated with stroke? |
| 70 | What are the causes for ischemic stroke? |
| 71 | What are the causes for hemorrhagic stroke? |
| 72 | What is atrial fibrillation? |
| 73 | Is smoking associated with stroke? |
| 74 | Is unhealthy diet associated with stroke? |
| 75 | What are the symptoms of a stroke? |
| 76 | What is a mini-stroke or transient ischemic attack (TIA)? |
| 77 | How is a stroke diagnosed? |
| 78 | What are the treatment options for stroke? |
| 79 | How common are strokes? |

**Table 2: Framework matrix showing attributes of responses to questions about general information about stroke.**

| **Participant** | **A : content** | **B : structure** | **C : tone** |
| --- | --- | --- | --- |
| 1: stroke survivor | It was purely factual, and I was asking, the question was asking for facts and I didn't feel it needed to be empathetic in any way. It was just what are the facts? And that's why I preferred it. I felt it was relevant to the question.  Yes, where, where, where there's no ambiguity, I don't think it needs to be empathetic. I think it just needs to be “these are the facts”. What are the risk factors of stroke? And here's your list. And this is it. Yes. So, I didn't feel there was any need for empathy. |  | I was thinking it was scary. |
| 2: stroke survivor |  | I think that one is quite blunt, so it just gives you a list of all the things which I think would be easier for people to understand |  |
| 3: stroke survivor |  |  |  |
| 4: stroke survivor | Didn't seem to contain as much detailed information |  |  |
| 5: carer | I found it easier to understand and I think it gave a little bit more detail. I just when the second one just said, you know TIA is a mini stroke and I just feel like that gives information |  |  |
| 6: stroke survivor |  |  |  |
| 7: stroke survivor | And also it used language like modifiable behaviours and it's kind of like, well, is everyone going to understand what that means?  Yeah, it used very kind of clinical language. |  | It felt quite blunt in the very first kind of paragraph, where it kind of went if you don't get treatment for a stroke, you can die.  Whereas the second one was much better at kind of explaining things in a more meaningful way. |
| 8: stroke survivor | Too much, too much medical information, which was too frightening for people who don't understand |  | Very clinical, not any personable information about it[...]I'm looking for something, maybe a wee bit of sugar-coated.  I found that very scary. |
| 9: stroke survivor | Too many facts, too much information. You don't need all that information. |  |  |
| 10: carer |  |  | just kind of the the language used seems a bit softer. |
| 11: stroke survivor |  |  |  |
| 12: stroke survivor |  |  |  |
| 13: stroke survivor | Lots of sort of medical terminology there, but you need to have the correct terms and I wouldn't fault it |  |  |
| 14: carer |  | It was the second one just read much more easily. It seemed much more on topic. And yeah, it ended up with me having better information, I thought |  |
| 15: stroke survivor |  |  |  |
| 16: stroke survivor | So I'll actually go to take my time and I've got to go back and have a look and maybe highlight and take out the bits that I need and I think. | I think, I think when I’d seen the first layout. And somebody's tried to generate AI answers in there. Then I've had a good look at that and went hold on a minute. This is, that may be relevant or may not be relevant, this is a bit easier to understand, especially someone like me. Not only getting old a bit but someone with uh…like my memories, how should I say it, I'm not, I'm not as fast. I'm not as reliable reading paragraphs like that.  I met the nurse and I spoke to the doctor and I spoke to the stroke people, they were all…brilliant information. Easy to understand which I need. And some, and the way some of the layouts were I found, they seem to go a bit to the side rather than stick with what happened, don’t they? |  |
| 17: stroke survivor |  | The layout was better in the second one. It had a bit of empathy and it was simple to read. Yeah, it was too waffly the first one. |  |
| 18: stroke survivor | Maybe just the fact that it used more kind of medical terms in this one yeah.  It's maybe it's kind of more kind of layman terms |  |  |

**Table 3: Framework matrix summarising attributes of responses to questions about health issues.**

| **Participant** | **A : content** | **B : structure** | **C : tone** |
| --- | --- | --- | --- |
| 1: stroke survivor | There were things, the thing that struck me was it talked about preeclampsia but didn't explain what it was. And it said at one point about menstrual issues, physical issues with menstruation, and it said go to GP who might recommend an occupational therapist, but it didn't say why. You know, I would have expected to say an occupational therapist who would be able to tell you about specific products or show you alternative methods or…it was sort of, oh, right, but what's the occupational therapist going to do? |  |  |
| 2: stroke survivor | I feel as if the first one had more kind of medical stuff in it because that one (B) was more kind of about different things. |  |  |
| 3: stroke survivor |  | It seemed to be more matter of fact and the way it was, it was laid out |  |
| 4: stroke survivor | It just kind of gave you facts what you need to know, rather than stuff like roundabout more detailed information |  |  |
| 5: carer |  | I feel like it was just very, like structured and laid out whereas this other one was a lot more like longer sentences |  |
| 6: stroke survivor |  |  | It seemed the first one stated facts. Just facts and somebody wrote that they need to be a bit more empathetic. |
| 7: stroke survivor | It kind of looked more formulaic. It it used more technical language. And it just kind of said things, said facts, words. It didn't kind of put any context or meaning on them.  With B it started off with that statement about informal carers, which kind of felt as though it understood the question a little bit better. | And both of them had quite long sentences. But I think with A, I found the sentences just a little bit harder to understand what it was trying to say. |  |
| 8: stroke survivor |  |  |  |
| 9: stroke survivor |  |  |  |
| 10: carer | I felt the second one had more, B had more information, so I think more of a medical professional would have wrote it. |  |  |
| 11: stroke survivor |  |  | the second response just seemed more comprehensive and more empathetic as if it came from a human. |
| 12: stroke survivor |  | It was too, not in depth, but it was more…I don't, I'm not really sure but just A seemed easier to understand and get to the point quicker than B did. |  |
| 13: stroke survivor |  |  |  |
| 14: carer |  |  |  |
| 15: stroke survivor | The only thing which isn't covered in these questions, a lot of it was just referrals on to other people, resources like contact the GP, contact 999, it doesn't, it's not a comprehensive answer. It's more like if you feel like this, do go talk to someone else, but that's not good.  It's not a deep dive. It's sort of an explanation. And then it basically refers you onwards to other information, but it's what it's doing and as well as giving you advice. You know, if you feel suicidal, contacting 999.  I prefer B. There was more information, including percentages in it. |  |  |
| 16: stroke survivor |  |  | It just seemed a little bit more robotic and a little bit more jargony. It didn't seem quite as approachable. |
| 17: stroke survivor |  |  | I know what they're saying is right but I haven't tried some of things, so I suppose I'm very unwilling. Maybe. |
| 18: stroke survivor |  | Well, I'm getting old and that and this information for me that was all very quick. And I don't concentrate the way that I used to concentrate before this happened to me, but I feel that B might have gave me a bit more time to actually see the causes and side effects. |  |

**Table 4: Framework matrix showing attributes of responses to questions about life after stroke.**

| **Participant** | **A : content** | **B : structure** | **C : tone** |
| --- | --- | --- | --- |
| 1: stroke survivor |  |  | I felt that B was more in layman's terms and also covered…I suppose, I thought it was more empathetic, I think that I would say with B it seemed to be more, it seemed to be less technical and more emotional, personal, yes. I much preferred B. |
| 2: stroke survivor |  |  |  |
| 3: stroke survivor |  |  |  |
| 4: stroke survivor | it was a more kind of generalised rather than specific to an individual person or a patient |  |  |
| 5: carer |  |  |  |
| 6: stroke survivor |  |  | Came across well the second one. It it just, again, it didn't just this is the fact, this the fact. It was more gentle. |
| 7: stroke survivor |  |  |  |
| 8: stroke survivor |  |  |  |
| 9: stroke survivor | It was lots of information. You might not need all the information. |  |  |
| 10: carer | Because it was a bit more information going, giving too much information. |  | It was less precise and a little bit colder. |
| 11: stroke survivor |  |  | it's a language used I felt B had a more empathetic tone to it |
| 12: stroke survivor |  |  |  |
| 13: stroke survivor |  |  |  |
| 14: carer |  |  |  |
| 15: stroke survivor | I'd say I prefer B because that's because it said that you know a care package will be put together by the team at the hospital, whereas A was like you need to contact your local social services department again, which is true, I don't know, but I prefer B I think. |  |  |
| 16: stroke survivor | It just didn't seem to be answering the question. It started off on completely the wrong track. It was answering what will my duties be as a carer for someone, rather than how do I find a carer. |  |  |
| 17: stroke survivor |  |  |  |
| 18: stroke survivor | That seems to be like a one size fits all.  There was a mixture in there that, I don’t know if it was just me, but if I had to read that back I would think that it was more to do with the person's looking after someone. Unless I've not picked that up right, I don't know.  Maybe I'm that, that, there may have been, there may have been information there that's not relevant to me, but again at the same time there may, there should possibly should have been information there that I’ve never seen. | They were quite long, both of them and to me it could have been for the carer or the victim, so to speak.  Yeah, I think I think if there was an AI then it would need to be separated into who you are. |  |

**Table 5: Framework matrix showing attributes of responses to questions about stroke recovery.**

| **Participant** | **A : content** | **B : structure** | **C : tone** |
| --- | --- | --- | --- |
| 1 : stroke survivor |  |  |  |
| 2 : stroke survivor | I didn't have that type of stroke, so I'm not sure if that's something that should happen or shouldn't, because I didn't he have any any of the stuff that's on that |  |  |
| 3 : stroke survivor |  |  | I think it was probably because it was very, what’s the word, unempathetic. Yeah. It was more sort of strict bullet points of boom, boom, boom. |
| 4 : stroke survivor | I felt B wasn't as kind of personalized, was more of a generic general response, not very specific for certain things. |  |  |
| 5 : carer |  |  |  |
| 6 : stroke survivor |  |  |  |
| 7 : stroke survivor |  |  |  |
| 8 : stroke survivor |  |  | It had a more of a human touch to it, just better understanding of what the issues are. |
| 9 : stroke survivor |  |  | Number one was more factual and not as empathetic as B. |
| 10 : carer | I thought A was more informative. For something so important regarding your driving, more information seem to be more factual and more important to be more factual. |  |  |
| 11 : stroke survivor |  |  | Again, it's just the tone of the language used. I feel like A was more empathetic, B was more clinical. |
| 12 : stroke survivor | I didn't actually prefer one above the other because some of the information in it I didn't agree with, so I don't, didn't really like any of them. |  |  |
| 13 : stroke survivor |  |  | I thought the second one felt more it come from, it was more emotional. Mostly the first one just seemed to give you the instructions as to the steps you would have to follow. The second one seemed to be a wee bit more empathetic. I don't know if that's AI playing a trick or not, but. |
| 14 : carer |  |  |  |
| 15 : stroke survivor | I mean, it's just a description of what people's jobs are. If I literally was typing that into a chatbot and that's what I got. I'll be like, OK, that explained to me the medical profession, not what I'm meant to do now and what I can do.  Again, I take it down a bit because it's acronyms in there. Again, looks more pointed at medical professionals rather than civilians.  It was very relevant, yeah. But again, like not the the information you want after you've had a stroke. So if there was a question, “would this be helpful?”, I don't think it was that helpful.  It looks like it's written by medical medic to sort of like try to explain the profession. And this is difference between an OT and this is a difference between a physio like grand, but that's not really what I want to know.  You'd need something like that if you're being discharged from hospital because your concern would be does my treatment stop? As I said, I'm just like being kicked out. So that's something that would explain that this is part of a sort of a phase and that's useful. | There was that typo type of mistake at the beginning and then there was a sentence that didn't make any sense in the first paragraph that would concern me. | But the way that's written, there's sort of describing things matter of fact, not from a patient point of view. It's too informative, not patient focused. |
| 16 : stroke survivor |  | A was really straightforward and down the line and B seemed to sort of meander around a bit more, was a bit more vague. But I definitely like A's presentation more |  |
| 17 : stroke survivor |  |  |  |
| 18 : stroke survivor | It was the way it was worded. I think I noticed in A, it didn't mention insurance plan, in B it did and I think there was something to do with the are you able to go back and try again about seeing someone? I never noticed that in A, but I'm sure I've seen it in the B. |  |  |

**Table 6: Framework matrix showing attributes of responses to questions about support after stroke.**

| **Participant** | **A : content** | **B : structure** | **C : tone** |
| --- | --- | --- | --- |
| 1: stroke survivor |  |  | It's a shame you haven't got overly empathetic. It was patronising in places.  It wasn't as patronising as A. Apart from the very first sentence where it says it was crucial to do all these things, and I didn't like that at all. It could be it is advisable. Other words, crucial is you have got to do this and it's just not for everybody at all.  Yeah, some people just, I'm not saying it's right, but some people…I think the information should be out there but tell somebody they've got to do it just eliminates personal choice. And the trouble with having a stroke is your personal choices are restricted anyway.  I felt, I felt it was a bit, again, it was a bit schoolteacher-ish. You know, you must do this and oh, do this because it's going to make you part of the community. And, you know, I mean, somebody like me, the thought of sitting there moaning to everybody else and having to listen to everybody else's problems doesn't appeal.  Yes, I don't like it when you’re told you have to do something. Yeah, fair enough if it's, you know, you're going to die if you don't have this injection. But even then there's an element of choice in that it's just say no, I'm not. Whereas it is crucial that you do this and it is essential. And oh, this wonderful sense of community. What if you don't want a sense of community? |
| 2: stroke survivor | This stuff's relevant, but I'd probably be against like things like YouTube and stuff like that because I did watch a couple of videos myself on YouTube to do exercises and when I spoke to my doctor who's a professional, he said that they don't work. |  |  |
| 3: stroke survivor |  |  |  |
| 4: stroke survivor | It gave an answer for across the board, it seemed to me like it included the answer for other countries, not just where. So it wasn't specifically helping for UK. It was too, too generalized. |  |  |
| 5: carer | It's a bit more like general, whereas like A was very specific. It lists all the different places which I don't know if I did like. So it's I almost didn't trust that it was given specifics anyway. |  |  |
| 6: stroke survivor |  |  | The information is there but it's just laid down in a kind of, this is it, this is it, this is...it doesn't say why. Doesn't seem to be forthcoming and saying "it would maybe help you to do this" or something. It just says go to that association, go to that association. And it's actually off-putting. If I read that, I'd probably go, oh ok, put it to the side. You probably never pick it up again. I think it would depend on the individual but I think after having a stroke, the biggest feeling is fear, fear of what is coming, fear. And then it never said that it can be hard to go to these groups. I found that very hard. |
| 7: stroke survivor |  | You know you can get this service in this town, this town, this town, this town, this town that, which isn't a very helpful way of giving that information and just felt AI generated. |  |
| 8: stroke survivor | A was just kind of like basic facts. B to me appeared to be more of the stuff that you worry about after you've had your stroke and explained a bit in a better way what benefits you can use and where else to go for help. It would give you more information other than what you can get off the Internet.  There was extra stuff in there to help. |  | When you become talking about finance, it's eh, it’s a matter of a fact stuff, isn't it?  B was, had more of a personable touch. |
| 9 : stroke survivor |  | Too much information. And it was a bit confusing. |  |
| 10: carer |  | Yes, number two was waffly, too much information, it wasn't relevant to be sort of a general thing, and the second one was very clear and concise. |  |
| 11: stroke survivor |  |  |  |
| 12: stroke survivor | Well, I think the first one was more geared towards stroke survivors, being part of a peer group and you probably can tell from my answers that I am a member of a peer group. And I think the second one, the AI one, was very general. |  |  |
| 13: stroke survivor | It just didn't seem to hold as much information. |  |  |
| 14: carer | Because more of the information on what you were looking for was given, yeah. |  |  |
| 15: stroke survivor | There was obviously was quite dense information, but that's because the support system is complicated. But it was easy to understand.  If you're in the UK, you want to know about the UK, but you know the thoughts are like it's a compilation of lots of information being pulled together in different countries. |  |  |
| 16: stroke survivor | And just the repeated slightly irrelevant details telling me about services, and Cardiff and Cardigan when I'm not in either of them. It was just a bit, B just felt a bit weird |  |  |
| 17: stroke survivor |  |  |  |
| 18: stroke survivor |  | I just felt B was easier to understand than A. |  |

**Table 7: Summary of participants' quotations to illustrate key themes.**

|  | **Themes** | | |
| --- | --- | --- | --- |
| **Topic** | **Content** | **Structure** | **Tone** |
| **General information** | Participants favoured detailed but factual and focused information. | Simple, direct list with clear, concise layout is easier to understand. | Excessive use of medical terms felt frightening and difficult to understand. Rephrasing information in plain language is beneficial. |
| **Health issues** | Context and detailed explanation of medical facts should be provided for all recommendations and signposting advice. | Short and clear sentences are preferred and easier to understand. | The tone of the response should be approachable, warm, empathetic and comprehensible instead of plain facts. |
| **Life after stroke** | Clear, specific information without excessive details is desired. The response should stay focused on the question, avoiding irrelevant information. | The response should be short and tailored towards the user category (stroke survivor, carer, family). | The tone of the response should be warm, empathetic and emotional. Plain language was favoured, while avoiding technical terms. |
| **Stroke recovery** | Participants felt that the responses should contain actionable specific advice, information with examples and explanations of medical procedures and terms. | Participants preferred clear, concise and structured layout. In contrast, bullet-point format was not well received. | Participants felt that a warm and empathetic person-centred tone is more relatable to the issues experienced after stroke. |
| **Stroke support** | Participants agreed that very localized information seemed unreliable. Country-specific rather than generalised or very localized responses seem to be optimal. | Participants agreed that the structure of the response should be clear, concise and easy to understand without being overly wordy. | Participants felt that responses may seem patronising and directive. Responses that motivate and suggest options while supporting choice and autonomy are preferred. |
